# On the survival of individuals diagnosed with type 2 diabetes mellitus in the United Kingdom: a retrospective matched cohort study

**DOI:** 10.1101/2021.12.13.21267749

**Authors:** Njabulo Ncube, Elena Kulinskaya, Nicholas Steel, Dmitry Pchejetski

**Author notes:** Corresponding Author: Njabulo Ncube, School of Computing, University of East Anglia, Norwich, Norfolk, United Kingdom, NR4 7TJ.

## Abstract

**Objective:** To estimate long-term hazards of all-cause mortality following a diagnosis of type 2 diabetes mellitus (T2DM) using electronic primary care data.

**Methodology:** Retrospective matched cohort study using electronic health records from THIN primary care database. The study included individuals born between 1930 and 1960, diagnosed with T2DM between 2000 and 2016 and aged 50-74 years and excluded those with pre-existing stroke, cancer, cognitive impairment, lower limb amputation or chronic kidney disease (CKD) stages 3 to 5. T2DM individuals were matched at diagnosis to at most 3 controls by age, gender and general practice (GP) and followed up to 1 January 2017. Time-varying hazards of all-cause mortality were then estimated using Gompertz-double-Cox model with frailty on GP, adjusting for medical history, socio-demographic and lifestyle factors.

**Results:** A total of 221 182 (57.6% Males, 30.8% T2DM) individuals were selected for the study of whom 29 618 (13.4%) died during follow-up. The adjusted mortality hazard of type 2 diabetes mellitus (T2DM) was estimated to be 1.21[1.12-1.3] and 1.52[1.44-1.6] among individuals diagnosed at 50-59 years and 60-74 years, respectively, compared to controls. Deprivation, obesity, smoking and comorbidities affected survival of cases and controls equally. Compared to the 1930-39 birth cohort, all-cause mortality hazards were reduced in the 1940-49 cohort, but increased at older ages in the 1950-60 birth cohort for both cases and controls.

**Conclusion:** T2DM is associated with raised all-cause mortality hazards which increase with age of diagnosis. These hazards associated with age at diagnosis are constant across all birth cohorts demonstrating a lack of progress over time in reducing the relative risks of all-cause mortality associated with T2DM. A further study that includes people born after 1960 is needed to fully understand the emerging higher mortality hazards among the younger birth cohorts.

**Significance of this Study:** *What is already known about this study?:*

- Diagnosis of type 2 diabetes mellitus (T2DM) is associated with increased all-cause mortality risk.

*What are the new findings?:*

- The increased all-cause mortality associated with T2DM was lower than previously reported.
- The all-cause mortality risk associated with T2DM increased with increasing age at diagnosis.
- The all-cause mortality risk associated with T2DM remained constant across all birth cohorts.
- The all-cause mortality risk is increased at later ages among the younger birth cohorts in individuals with and without T2DM.

*How might these results change the focus of research or clinical practice?:*

- Further research is needed on individuals born after 1960 to explore the increased all-cause mortality hazards in recent birth cohorts.

## INTRODUCTION

The global prevalence of diabetes mellitus (DM) is on the increase due to an ageing population and harmful lifestyles,[1-3]. About 463 million individuals worldwide were reported to be diagnosed with DM in 2019 and about 1.5 million deaths were caused by diabetes mellitus (DM) in the same year,[4, 5]. Since 2016 it has ranked among the top ten causes of death. In the UK, a new case of DM is diagnosed every two minutes and at least 4.8 million people were thought to be diabetic in 2019, of whom 20% had undiagnosed diabetes,[2]. The number of deaths caused by DM increased by 31% between 2013 and 2019,[6]. The cost of treating diabetes in the UK was 10% of the total prescribing cost in the year 2018/2019 (£10 billion).

Uncontrolled blood glucose level can cause complications such as cardiovascular diseases (CVD), eye, renal and neurological diseases and premature death. In the UK every week diabetic individuals have 175 amputations, 687 strokes, 530 myocardial infarctions MI and 2 000 episodes of heart failures HF while 30 lose their sight and 700 suffer premature death,[2]. In The Health Improvement Network (THIN) data, 91.1% of diabetics had type 2 diabetes mellitus (T2DM),[7]. T2DM is a chronic disease that occurs due to the pancreas failing to produce enough insulin or the body failing to use its insulin,[8]. It is an insidious disease that can continue for several years without being diagnosed. In the UK, in people of white origin the average age of diagnosis is 58 years while in Blacks and Asians it is 48 years and 46 years, respectively,[9]. Age, family history, ethnicity, sedentary lifestyle, smoking, hypertension (HTN), poor mental health and obesity have been identified as the main risk factors for T2DM,[2, 10, 11]. Obesity accounted for 80-85% of the risk of developing T2DM while being BAME increased the risk by 2 to 4 times,[2, 9]. T2DM individuals in the UK, were 50% more likely to die prematurely,[2].

Several previous studies have reported that people with T2DM experienced more than double the risk of mortality compared to people without T2DM,[12-15]. Mortality rates among individuals with T2DM are considerably higher than in the general population and the years of life lost due to T2DM were estimated to be 3 to 5 years in the white population aged 65 years and above,[16, 17].

Previous studies on T2DM were mainly pharmacovigilance based. The few and limited observational cohort matched studies on all-cause mortality adjusted for age, sex, smoking status, BMI or deprivation at most and had short follow-up periods (7.8 years),[14, 15]. The only study with a long follow-up, 20 years, had a smaller study population (13 000) compared to this study,[12] while another study included all diabetic individuals in their study,[13]. In addition, no previous studies estimated the effect of age of diagnosis and birth cohort on all-cause mortality. The study population in some previous studies constituted older T2DM individuals compared to non-diabetics which maybe a source of bias. This study aimed to estimate the survival prospects of individuals after diagnosis of T2DM in comparison to nondiabetic controls using data from UK primary care electronic health records (EHR) adjusting for more covariates (including lifestyle, socio-demographic factors and medical history) than previous studies.

## STUDY DESIGN

### Data Source

The study used The Health Improvement Network (THIN), a UK primary care database which in 2017 included medical records for 15.6 million individuals collected from 711 registered GPs. Most registered GPs contributed data for at least 20 years. Actively registered individuals in THIN represent about 6% of the UK population,[18]. Several studies have found THIN to be broadly representative of the UK population and hence suitable for inferential studies,[19-23]. This study was approved by THIN Scientific Review Committee (SRC) with approval number 16THIN095.

### Selection Criteria

Individuals born between 1930 and 1960 inclusive, diagnosed with T2DM between 2000 and 2016 inclusive and aged 50 years and above at diagnosis were selected. Of these, individuals with prior stroke, cognitive impairment including dementia, cancer, chronic kidney disease (CKD) stages 3-5 and lower limb amputations were excluded, as well as individuals with severe heart failure (HF), myocardial infarction (MI), atrial fibrillation (AF) and peripheral vascular disease (PVD). In addition, individuals with data gaps before conversion to Vision (a recording system used by general practices in THIN), no medical record within 10 years before diagnosis or flagged as not acceptable for research by internal THIN quality control were also excluded. Eligible T2DM individuals were then matched to at most 3 non-diabetics using similar selection criteria by age, GP and gender. Data was extracted using medical Read codes for diagnoses and the BNF drug codes. The Read codes were compiled from both the Clinical Codes website,[24] and by querying the THIN database. The Read codes and the extraction process are shown in *Sections A and B of the Supplementary*, respectively.

### Study Variables and Categorisation

The selected individuals were categorised into 3 birth cohorts (1930-1939, 1940-1949 and 1950-1960). The study used the quintiles of the patient postcode-based Townsend Deprivation Index (TDI) as the socio-economic status variable,[25]. TDI quintiles classify the population into 5 categories, with 1 associated with areas of least deprivation and 5 associated with the most deprived areas (further description is given in *Section C*.*4 of the Supplementary*). Pre-existing medical conditions which included HF, MI, AF and PVD were binary coded (No: -no diagnosis or Yes: -with diagnosis) while hypertension (HTN) and hypercholesterolemia (HCL) were both coded at three levels as “no diagnosis”, “treated” or “untreated”. The other variables included were gender, BMI (classified as “normal” [18.5≤ BMI*<*25], “overweight” [25≤ BMI*<*30] or “obese” [BMI≥30]), smoking status (classified as non-smoker, former smoker and smoker) and age at entry or diagnosis (categorised as 50-59 or 60-74 years old). Details of the derivation of variables and their coding are provided in *Section C of the Supplementary*.

### Statistical Analysis

#### Survival Model

The Cox regression model is a widely used survival analysis method. However, it requires a strong assumption of proportional hazard (PH) which often gets violated in empirical data. Instead, we used a generalized Gompertz-double-Cox survival model with frailty, introduced as Gompertz parametric model in Begun et al.,[26]. We refer to this model as a Gompertz-double-Cox as it is an extension of the Cox model that also includes a shape sub-model reparametrised as in a Cox model. This model uses the Gompertz baseline hazard function, Gamma-distributed random effect of the GP practice (frailty) and allows to model covariates’ effects on both shape and scale parameters. Prior to choosing this model, we ascertained that the hazards followed the Gompertz distribution (Figure S3). Covariates with significant shape effects present time-varying hazards. The model is briefly described in more detail in *Section C of the Supplementary*. The analysis was performed using RStudio 1.3.5 and R 4.0.1 software.

#### Missing Values

Missing values in covariates were handled using the joint model multiple imputation method [27, 28]. The joint modelling (JOMO) package in R was used, using the imputation model obtained from the complete-case data,[27]. Distributions of the imputed variables were compared to the complete-case data. Fifteen imputed data sets were created. The substantive model obtained from the complete-case analysis was then fitted on each of these datasets. For inference, the results were combined using the Rubin’s rules,[28].

#### Model Validation

The final model was selected using the backward AIC-based elimination with 5% and 1% significance levels applied for main effects and interactions, respectively. Coefficient of concordance,[29] was used to ascertain the model’s goodness-of-fit. For consistency, complete case model estimates were compared to the imputed data estimates. Results from similar previous studies were used for external validation.

## RESULTS

### Study population characteristics

Study population included a total of 221,182 (57.6% males, 30.8% T2DM) individuals selected from THIN (Table 1). The total mortality rate was 15.81 per 1000 person-years. The average age at diagnosis of T2DM was slightly above 61 years across the entry period.

**Table 1:** Study Population.

| Description | Entry Cohort |  |  |  |  |  |
| --- | --- | --- | --- | --- | --- | --- |
|  | 2000-2004 |  | 2005-2009 |  | 2010-2016 |  |
|  | T2DM | Diabetic-free | T2DM | Diabetic-free | T2DM | Diabetic-free |
| Numbers | 25 333 | 64 334 | 27 403 | 61 155 | 15 463 | 27 494 |
| Birth Cohort (%) |  |  |  |  |  |  |
| 1930-1939 | 28.10 | 71.90 | 30.20 | 69.80 | 34.70 | 65.30 |
| 1940-1949 | 28.30 | 71.70 | 30.70 | 69.30 | 35.50 | 64.50 |
| 1950-1960 | 28.90 | 71.10 | 30.90 | 69.10 | 36.50 | 63.50 |
| Males (%) | 58.70 | 56.80 | 59.70 | 56.50 | 60.30 | 57.00 |
| Deaths | 4 425 | 11 891 | 3 150 | 7 308 | 996 | 1 848 |
| Age (%) |  |  |  |  |  |  |
| 50-59 | 42.1 | 41.91 | 41.99 | 41.09 | 40.21 | 39.86 |
| 60-74 | 57.9 | 58.09 | 58.01 | 58.91 | 59.79 | 60.14 |
| mean (in years) | 61.22 | 61.26 | 61.36 | 61.51 | 61.65 | 61.64 |
| standard deviation | 6.44 | 6.44 | 6.75 | 6.75 | 6.31 | 6.36 |
| Mean Follow-up Time (Years) | 10.95 | 11.14 | 7.65 | 7.85 | 3.98 | 4.17 |
| Smoking Status (%) |  |  |  |  |  |  |
| Nonsmoker | 45.05 | 45.36 | 45.43 | 49.82 | 45.02 | 51.01 |
| Former | 27.60 | 18.12 | 34.66 | 26.04 | 35.10 | 27.70 |
| Smoker | 20.56 | 21.81 | 19.26 | 21.10 | 19.81 | 20.61 |
| Missing | 6.79 | 14.71 | 0.65 | 12.04 | 0.06 | 0.70 |
| Townsend Index (%) |  |  |  |  |  |  |
| Less Deprived | 21.63 | 27.17 | 21.94 | 26.89 | 20.74 | 26.69 |
| 2 | 20.36 | 22.93 | 20.38 | 22.93 | 20.09 | 22.34 |
| 3 | 20.19 | 18.78 | 20.1 | 19.07 | 19.88 | 19.10 |
| 4 | 18.35 | 15.36 | 18.00 | 15.01 | 19.04 | 15.02 |
| Most Deprived | 12.88 | 9.70 | 12.92 | 9.59 | 13.06 | 9.56 |
| Missing | 6.59 | 5.06 | 6.66 | 6.51 | 7.19 | 7.29 |
| BMI (%) |  |  |  |  |  |  |
| Normal Weight | 8.79 | 30.71 | 8.41 | 31.14 | 7.87 | 30.79 |
| Overweight | 30.49 | 31.31 | 29.36 | 35.46 | 27.85 | 37.23 |
| Obese | 45.12 | 13.02 | 54.44 | 18.36 | 58.84 | 21.99 |
| Missing | 15.61 | 24.95 | 7.79 | 15.04 | 5.44 | 9.99 |
| mean ($kg/m^2$) | 31.25 | 26.38 | 32.04 | 26.92 | 32.65 | 27.31 |
| standard deviation | 5.85 | 4.49 | 6.21 | 4.82 | 6.57 | 5.07 |
| HTN (%) |  |  |  |  |  |  |
| No Diagnosis | 23.33 | 46.75 | 23.54 | 45.34 | 27.99 | 48.12 |
| Treated | 67.76 | 32.98 | 63.34 | 32.31 | 52.6 | 27.56 |
| Untreated | 8.92 | 20.26 | 13.13 | 22.35 | 19.42 | 24.31 |
| HCL (%) |  |  |  |  |  |  |
| No Diagnosis | 77.51 | 90.21 | 58.50 | 74.39 | 50.04 | 61.84 |
| Treated | 21.79 | 7.55 | 39.55 | 15.73 | 44.32 | 18.05 |
| Untreated | 0.71 | 2.24 | 1.95 | 9.88 | 5.65 | 20.11 |
| Other Medical Conditions (%) |  |  |  |  |  |  |
| AF | 10.96 | 9.71 | 8.06 | 7.70 | 4.99 | 5.42 |
| HF | 11.67 | 6.85 | 11.24 | 6.99 | 9.58 | 6.25 |
| MI | 16.5 | 8.15 | 14.39 | 7.95 | 12.42 | 6.78 |
| PVD | 11.66 | 11.65 | 11.9 | 12.34 | 12.53 | 12.75 |

The prevalence of smokers, former smokers and non-smokers was 20.9%, 25.8% and 47.2%, respectively. The prevalence of former smokers increased by the year of entry in individuals both with and without T2DM. Forty-seven percent of the study population were from more affluent areas (TDI=1 or 2). In total, considerably more T2DM individuals were obese (52% versus 16% controls) than normal weight (8.4% vs 30.9%, respectively). The prevalence of hypertension (HTN) was 75.5% in T2DM compared to 53.6%, in controls, with more T2DM individuals having treated HTN than non-diabetics. HCL was also more prevalent in T2DM compared to controls. AF, HF, MI and PVD, all had prevalence less than 15% in both the T2DM and non-diabetic individuals.

Table 2 shows that the 1930-1939 birth cohort included only individuals aged 60-74 years at entry, while the 1950-1960 birth cohort was mainly made up of individuals aged 50-59 years (90.96%). It is important to note this age distribution as it helps in the interpretation of the estimated survival in Section 3.2.

**Table 2:**
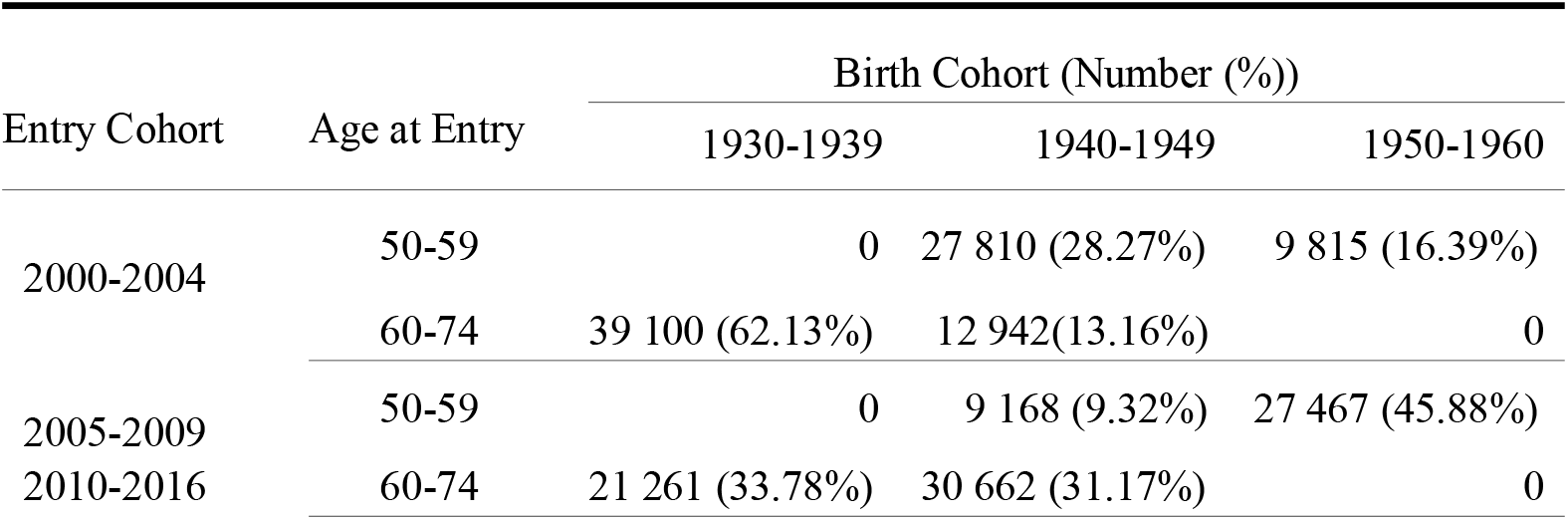
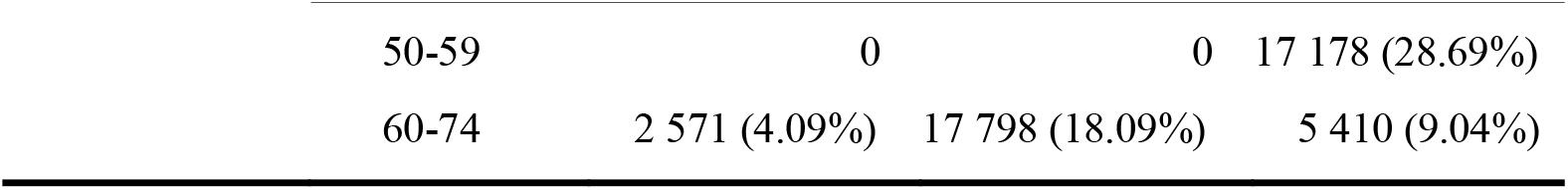
Study Population by age group, birth and entry cohorts

| Entry Cohort | Age at Entry | Birth Cohort (Number (%)) |  |  |
| --- | --- | --- | --- | --- |
|  |  | 1930-1939 | 1940-1949 | 1950-1960 |
| 2000-2004 | 50-59 | 0 | 27 810 (28.27%) | 9 815 (16.39%) |
|  | 60-74 | 39 100 (62.13%) | 12 942 (13.16%) | 0 |
| 2005-2009 | 50-59 | 0 | 9 168 (9.32%) | 27 467 (45.88%) |
| 2010-2016 | 60-74 | 21 261 (33.78%) | 30 662 (31.17%) | 0 |

|  |  |  |  |
| --- | --- | --- | --- |
| 50-59 | 0 | 0 | 17 178 (28.69%) |
| 60-74 | 2 571 (4.09%) | 17 798 (18.09%) | 5 410 (9.04%) |

Three variables had missing values, smoking status (7%), BMI (5%) and TDI (4%). The missing values for the smoking status and BMI decreased by entry year cohort and for T2DM individuals. Missing values increased for TDI by entry year cohort.

### Results of the survival modelling

The final model on imputed data was adjusted for birth cohort in 10-year intervals from 1930 to 1960, T2DM indicator with age group at diagnosis (50-59 or 60-74), gender, smoking status, TDI, AF, HF, HCL, HTN, MI, PVD and BMI group and significant interactions (Table 3 and Figure 1). The model had a concordance correlation of 0.75 (0.002 standard deviation) which was 1 percentage point above that of the similar complete cases model (0.74 (0.002).

**Table 3:**
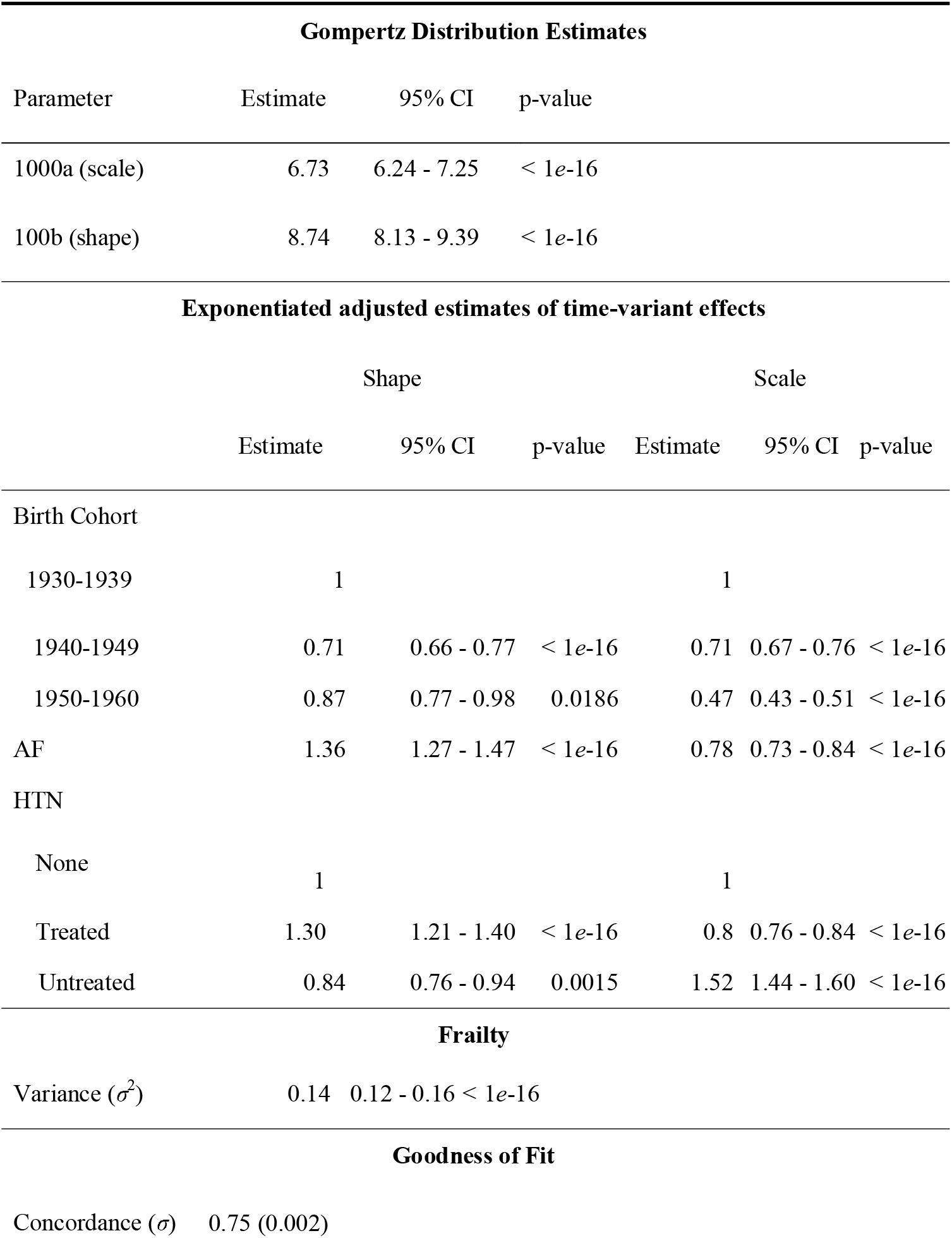
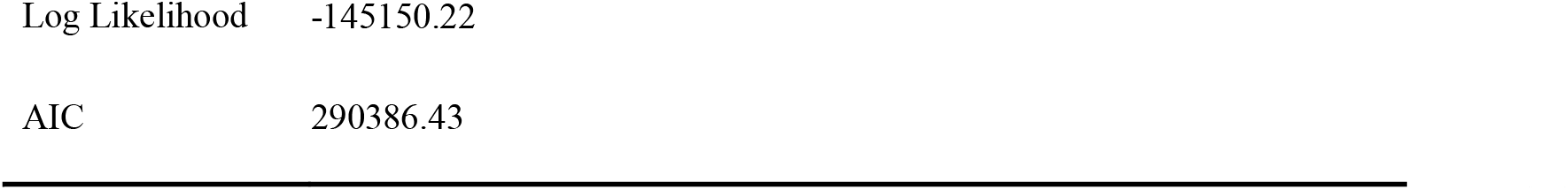
Estimates of adjusted scale, shape and frailty parameters and model goodness-of-fit statistics^†^

**Figure 1:**
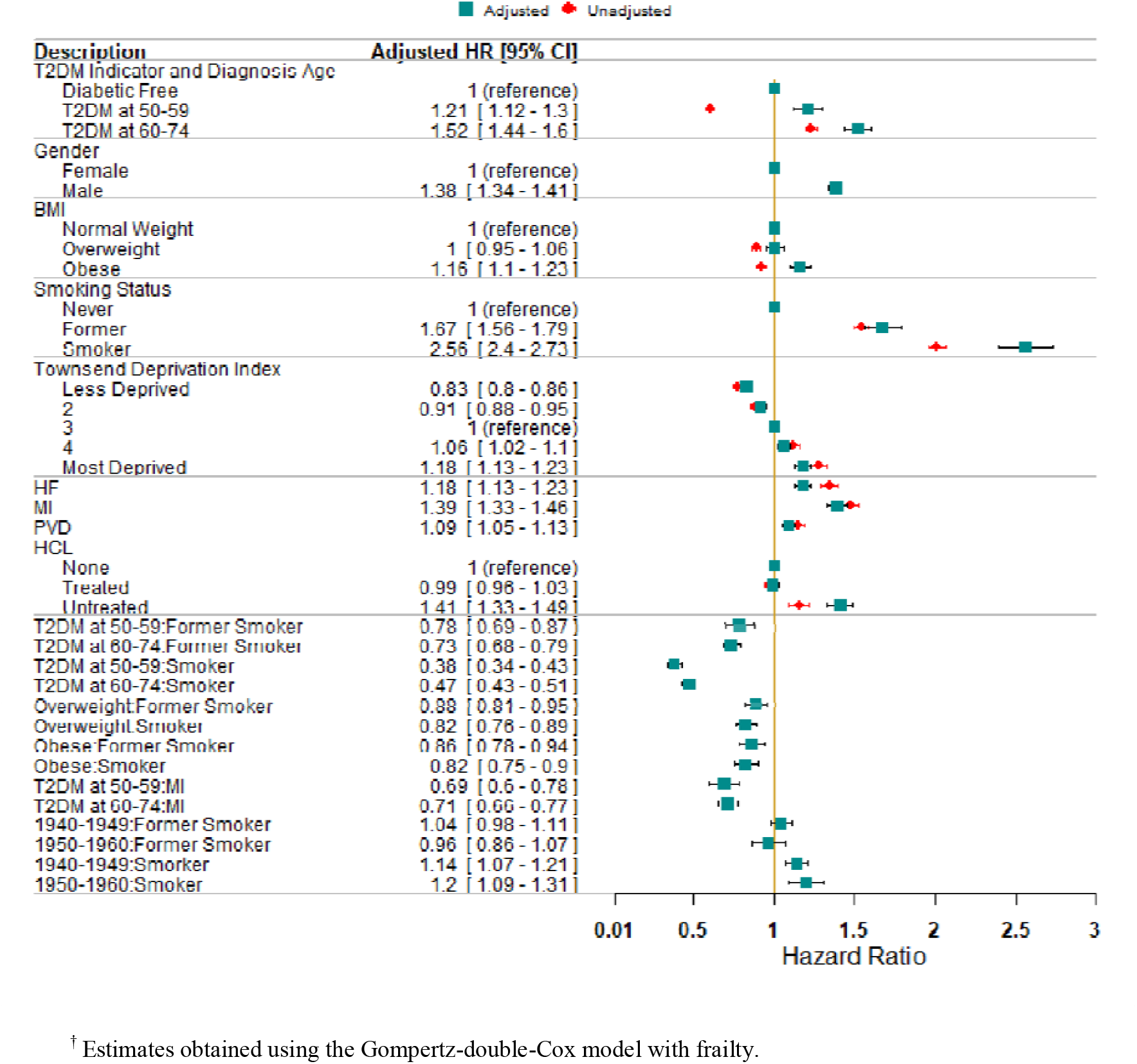
Adjusted and unadjusted HRs of all-cause mortality for covariates with significant time-invariant effects.

The baseline Gompertz distribution scale and shape parameters were estimated to be 0.0067 with a 95% confidence interval (CI) of [0.0062-0.0072] and 0.087 [0.0810.094], respectively. These parameters corresponded to the baseline hazard function for non-diabetic females aged 60-74 years at entry, born between 1930 and 1939, from a medium deprived area (TDI=3), non-smokers, of normal weight, with no AF, HF, MI, PVD, HTN and HCL. As both baseline scale and shape estimates are small, the baseline population had comparatively low mortality hazards. Three variables, namely the birth cohort, AF and HTN had significant time-varying effects on mortality.

Mortality hazards of variables with time-invariant effects compared to the baseline are depicted in Figure 1. The overall all-cause mortality hazard associated with T2DM is 1.38 [1.32-1.44] compared to non-diabetics. The results show an increased mortality risk with increase in age at T2DM diagnosis. A diagnosis between 60 to 74 years of age increased mortality risk by 25.6% percentage points compared to a diagnosis between 50 to 59 years of age (HR: 1.52 [1.44-1.6] versus 1.21[1.12-1.3]). These hazards were constant across all birth cohorts. Males had 38% increased hazard of mortality compared to females. Obesity was associated with a 1.16 [1.1-1.23] HR while being “overweight” was not statistically significantly different from being of normal weight. The effect of smoking on mortality increased by birth cohort.

Individuals from the most deprived areas (TDI=5) had a higher risk of mortality (HR of 1.18[1.13-1.23]) when compared to those from areas of medium deprivation (TDI=3), whereas the least deprived areas had a reduced hazard (HR of 0.83 [0.800.86]]). Having a pre-existing MI or PVD increased mortality risk by 39% and 9% compared to those with no diagnosis of MI or PVD, respectively. Untreated HCL had a statistically significant 41% mortality hazard increase, while the mortality hazards of individuals with treated HCL did not differ significantly from those without HCL. The mortality hazard associated with birth cohort, AF or treated HTN over time can be fully understood graphically as shown in Figure 2.

**Figure 2:**
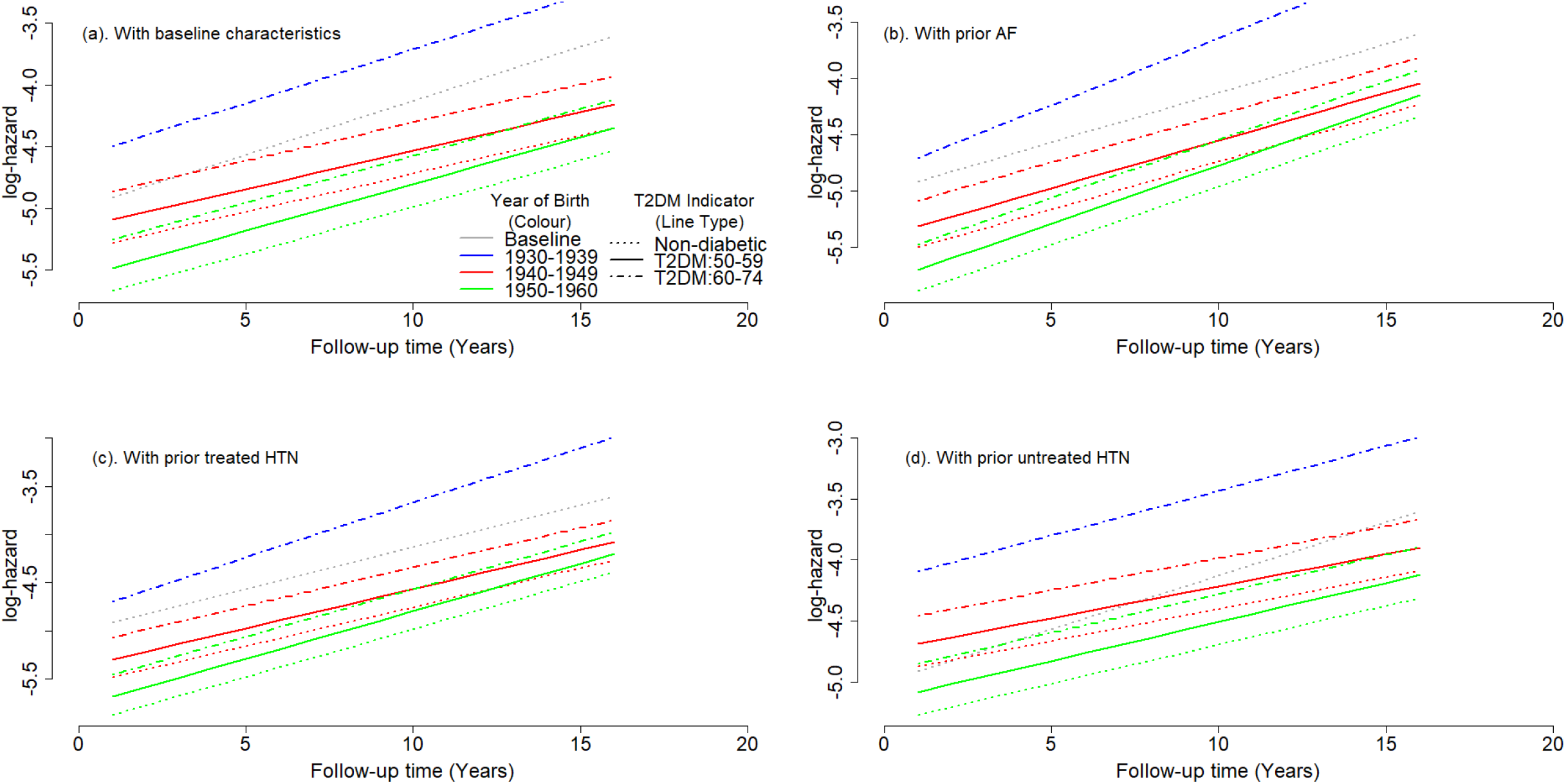
Comparison of the cumulative hazards of all-cause mortality of the T2DM individuals diagnosed at 60-69 years of age and their matched controls depicting time-varying mortality risk effects.

The 1930-1939 and 1940-1949 birth cohorts had higher mortality hazards than the 1950-1960 birth cohort at study entry. However, the 1950-1960 birth cohort showed higher mortality hazards at older ages than the 1940-1949 birth cohort. This indicates lower mortality improvements over time for both individuals with and without T2DM in the 1950-1960 birth cohort, *see* Figure 2a.

Hypertension and AF increased mortality risk compared to the baseline population as shown in Figures 2c to 2d. Taking antihypertensive drugs reduced the mortality hazard associated with HTN (Figures 2c and 2d).

## DISCUSSION

This large retrospective matched cohort study found, after adjusting for socio-demographic, lifestyle and medical factors, that individuals with T2DM experienced higher mortality hazards than non-diabetics and these hazards increased with age at diagnosis. The hazards associated with the age at diagnosis of T2DM remained constant across all birth cohorts. The prescription of antihypertensive or anti-hypercholesterolaemia drugs was associated with improved survival compared to no prescription for the relevant conditions.

T2DM has previously been reported to be associated with an increased mortality HR from 1.5 to above 2 compared to non-diabetics,[12, 14, 15]. These estimated HRs from previous studies were higher than HRs estimated in this study and did not consider the hazards associated with age at diagnosis. By combining all T2DM individuals regardless of age of diagnosis, previous studies overestimated all-cause mortality hazards associated with diagnosis of T2DM at younger ages. Lind et.al,[13] found a declining trend in mortality relative risk among T2DM individuals compared to non-diabetics between 1996 and 2009. However, this study found that the hazard ratios of all-cause mortality associated with T2DM remained constant across all birth cohorts.

The decline in all-cause mortality hazards among T2DM as reported by Lind et.al,[13] has previously been attributed to medical advancement, improved T2DM guidelines and management,[30-35]. Another study by Lim et.al,[36] found mixed outcomes and suggested that mortality improvement was dependent on the treatment. However, by backward elimination, the effects of antidiabetic drugs and treatment intensification on all-cause mortality were found to be not statistically significant and hence not included in the final model of this study.

There have been a large number of studies on T2DM, but its effect on all-cause mortality compared to non-diabetics has not been extensively studied in the UK and no study has estimated the effect of age at diagnosis. Existing literature has mainly been on T2DM pharmacosurveillance. The few relevant studies adjusted for at most 3 variables (age, gender, smoking status or entry year),[13-15]. Exclusion of variables such as the birth year, whether grouped or not, averages the study population survival prospects across all years which can increase relative risk bias and confounding. By adjusting for a considerable number of risk factors, this large study minimised biases in estimation of main effects and confounding. To minimise selection and information bias and bias by indication age at entry, GP and gender were used to match T2DM patents to non-diabetics. Furthermore, GP was used as a latent (frailty) variable to account for risk homogeneity among individuals who shared the same risks based on medical services received. Imputed data was validated against the complete data and no differential outcome was found as a result of imputation. However, though the study included several statistically significant variables, these were not exhaustive. Hence, there may be potential residual confounding due to unavailable data such as changes in therapy or lifestyle, severity of smoking and other unrecorded variables.

This study also validated previous findings on the benefits of anti-hypertensive treatment compared to no treatment,[37-41]. However, the cited studies had a short follow-up time and HTN was modelled as a time-invariant factor. As this study has shown, the hazard of HTN on all-cause mortality had a significant shape effect and hence should be modelled as time-variant. The study also found that the hazards due to smoking increased in the later birth cohorts. An important finding of this study is the increase in all-cause mortality hazards at later ages in the 1950-60 birth cohort for individuals with or without T2DM. Compared to the 1930-39 birth cohort, all-cause mortality hazards reduced in 1940-49 cohort, but increased at older ages in the 1950-60 birth cohort for both cases and controls. This finding is in line with a recent study by Rashid et.al,[42] that found that the number of middle-layer super output areas (MSOA) in England with a decline in life expectancy in women increased by 262% in 2014-2019 compared to 2010-2014, out of the total of 6791 MSOAs. For men, 11.5% of MSOAs had a decline in life expectancy in 2014-2019. Though the study,[42] was based on the England population, England has a respectable 84.2% of the UK population. In addition, their findings indicated that the poor changes in mortality in the UK, began in the decade before Covid-19 pandemic. This study has identified sources of this poor change in mortality, for example the 1950-1960 birth cohort and an increasing effect of smoking by birth cohort.

In comparison to the complete case model, which included 69.6% of study population and 66.2% of deaths, the imputed model had a concordance of 0.75 which was one percentage point higher (for complete case model, *see Table S2 and Figure S2 in the Supplementary*). The imputed model provided similar HRs with reduced variance and higher significance compared to the complete case model.

The Cox model is the standard method of survival analysis used in assessing the risk of mortality. By its very nature, the Cox model has a strong assumption on PH. The model by Begun et.al,[26] is a generalisation of the Cox model that allows for modelling of the baseline hazards shape parameter to adjust for time-variant variables while retaining the Cox model structure. This model has a better statistical power in case of time-variant hazards. Study limitations include the lack of ethnicity and antidiabetic drugs in our models though they are important variables. Ethnicity was excluded due to high percentage of missing values in THIN. Another important variable excluded due to high number of missing values was HbA_1*c*_.

In conclusion, the HRs associated with T2DM were somewhat lower than previously reported, at 1.21-1.52. However, the hazards were constant across all birth cohorts demonstrating a lack of progress in reducing relative risks of mortality associated with T2DM over time. Pre-existing medical conditions and in particular untreated HTN and smoking increased the mortality hazards and their effects increased in later birth cohorts for both T2DM and non-diabetics. In addition, this study has found that the mortality hazards were higher at older ages in the younger birth cohort.

The poor mortality experience in the 1950-1960 birth cohort, merits further research on individuals born after 1960 to explore the increased all-cause mortality hazards in recent birth cohorts.

## Data Availability

All data produced in the present study are available upon reasonable request to the authors.

## Contributors

All authors have contributed sufficiently to the conception or design of the work and to, interpretation, drafting the work or critically revising it and approved the work presented in the manuscript to be published and all take public responsibility for the content. EK and NN were responsible for the acquisition and analysis of the data. NN was responsible for the data extraction.

## Ethical Approval

This study was approved by THIN Scientific Review Committee (SRC) with approval number 16THIN095.

## Acknowledgments

The authors would like to thank Dr. Alexander Begun for providing R package implementing his model and the support he gave in customising the package to suit the study design and analysis.

