## Supplementary for "On the survival of individuals diagnosed with type 2 diabetes mellitus in the United Kingdom: a retrospective matched cohort study"

### A THIN Medical Codes

Type 2 diabetes mellitus (T2DM) readcodes used in this study were collected from the [Clinical Codes.](http://www.clinicalcodes.org/) Readcodes collected from this website included other diabetes mellitus (DM) types which were excluded from the study as per medical advice by the project medical professionals.

Table S1: T2DM Readcodes used in the study.

| Readcode | | | Description | | | | | Source | | Included | |
| --- | --- | --- | --- | --- | --- | --- | --- | --- | --- | --- | --- |
| C11y000 | | | Steroid induced diabetes | | | | | Clinicalcodes | | YES | |
| C314.11 | | | Renal diabetes | | | | | Clinicalcodes | | YES | |
| 9N4p.00 | | | Did not attend diabetic retinopathy clinic | | | | | Clinicalcodes | | YES | |
| 66At111 | | | Type 2 diabetic dietary review | | | | | Clinicalcodes | | YES | |
| C109K00 | | | Hyperosmolar non-ketotic state in type 2 diabetes mellitus | | | | | Clinicalcodes | | YES | |
| C109J12 | | | Insulin treated Type II diabetes mellitus | | | | | Clinicalcodes | | YES | |
| C109J11 | | | Insulin treated non-insulin dependent diabetes mellitus | | | | | Clinicalcodes | | YES | |
| C109J00 | | | Insulin treated Type 2 diabetes mellitus | | | | | Clinicalcodes | | YES | |
| C109H12 | | | Type 2 diabetes mellitus with neuropathic arthropathy | | | | | Clinicalcodes | | YES | |
| C109H11 | | | Type II diabetes mellitus with neuropathic arthropathy | | | | | Clinicalcodes | | YES | |
| C109H00 | | | Non-insulin dependent d m with neuropathic arthropathy | | | | | Clinicalcodes | | YES | |
| C109G11 | | | Type II diabetes mellitus with arthropathy | | | | | Clinicalcodes | | YES | |
| C109G00 | | | Non-insulin dependent diabetes mellitus with arthropathy | | | | | Clinicalcodes | | YES | |
| C109F12 | | | Type 2 diabetes mellitus with peripheral angiopathy | | | | | Clinicalcodes | | YES | |
| C109F11 | | | Type II diabetes mellitus with peripheral angiopathy | | | | | Clinicalcodes | | YES | |
| C109F00 | | | Non-insulin-dependent d m with peripheral angiopath | | | | | Clinicalcodes | | YES | |
| C109E12 | | | Type 2 diabetes mellitus with diabetic cataract | | | | | Clinicalcodes | | YES | |
| 66AH200 | | | Conversion to insulin by diabetes specialist nurse | | | | | Clinicalcodes | | YES | |
| C109E11 | | | Type II diabetes mellitus with diabetic cataract | | | | | Clinicalcodes | | YES | |
| C109E00 | | | Non-insulin depend diabetes mellitus with diabetic cataract | | | | | Clinicalcodes | | YES | |
| C109D12 | | | Type 2 diabetes mellitus with hypoglycaemic coma | | | | | Clinicalcodes | | YES | |
| C109G12 | | | Type 2 diabetes mellitus with arthropathy | | | | | Clinicalcodes | | YES | |
| U602317 | | | [X] Adverse reaction to glipzide | | | | | Clinicalcodes | | YES | |
| 66At100 | | | Type II diabetic dietary review | | | | | Clinicalcodes | | YES | |
| ZC2C800 | | | Dietary advice for diabetes mellitus | | | | | Clinicalcodes | | YES | |
| ZC2CA00 | | | Dietary advice for type II diabetes | | | | | Clinicalcodes | | YES | |
| ZRB5.00 | | | Diabetes treatment satisfaction questionnaire | | | | | Clinicalcodes | | YES | |
| ZRB5.11 | | | DTSQ - Diabetes treatment satisfaction questionnaire | | | | | Clinicalcodes | | YES | |
| ZRB6.00 | | | Diabetes wellbeing questionnaire | | | | | Clinicalcodes | | YES | |
| C10FC00 | | | Type 2 diabetes mellitus with nephropathy | | | | | Clinicalcodes | | YES | |
| C10FH11 | | | Type II diabetes mellitus with neuropathic arthropathy | | | | | Clinicalcodes | | YES | |
| C10FH00 | | | Type 2 diabetes mellitus with neuropathic arthropathy | | | | | Clinicalcodes | | YES | |
|  | C10FG11 | | | | Type II diabetes mellitus with arthropathy | | | Clinicalcodes | | | YES |
|  | C10FG00 | | | | Type 2 diabetes mellitus with arthropathy | | | Clinicalcodes | | | YES |
|  | C10FF11 | | | | Type II diabetes mellitus with peripheral angiopathy | | | Clinicalcodes | | | YES |
|  | C10FF00 | | | | Type 2 diabetes mellitus with peripheral angiopathy | | | Clinicalcodes | | | YES |
|  | C10FE11 | | | | Type II diabetes mellitus with diabetic cataract | | | Clinicalcodes | | | YES |
|  | C10FE00 | | | | Type 2 diabetes mellitus with diabetic cataract | | | Clinicalcodes | | | YES |
|  | C10FD11 | | | | Type II diabetes mellitus with hypoglycaemic coma | | | Clinicalcodes | | | YES |
|  | C10FC11 | | | | Type II diabetes mellitus with nephropathy | | | Clinicalcodes | | | YES |
|  | C10FK00 | | | | Hyperosmolar non-ketotic state in type 2 diabetes mellitus | | | Clinicalcodes | | | YES |
|  | C10FB11 | | | | Type II diabetes mellitus with polyneuropathy | | | Clinicalcodes | | | YES |
|  | C10FB00 | | | | Type 2 diabetes mellitus with polyneuropathy | | | Clinicalcodes | | | YES |
|  | C10FA11 | | | | Type II diabetes mellitus with mononeuropathy | | | Clinicalcodes | | | YES |
|  | C10FA00 | | | | Type 2 diabetes mellitus with mononeuropathy | | | Clinicalcodes | | | YES |
|  | C10F911 | | | | Type II diabetes mellitus without complication | | | Clinicalcodes | | | YES |
|  | C10F900 | | | | Type 2 diabetes mellitus without complication | | | Clinicalcodes | | | YES |
|  | C10F711 | | | | Type II diabetes mellitus - poor control | | | Clinicalcodes | | | YES |
|  | C10F700 | | | | Type 2 diabetes mellitus - poor control | | | Clinicalcodes | | | YES |
|  | C10F611 | | | | Type II diabetes mellitus with retinopathy | | | Clinicalcodes | | | YES |
|  | C10FD00 | | | | Type 2 diabetes mellitus with hypoglycaemic coma | | | Clinicalcodes | | | YES |
|  | C10FR00 | | | | Type 2 diabetes mellitus with gastroparesis | | | Clinicalcodes | | | YES |
|  | 13AC.00 | | | | Diabetic weight reducing diet | | | Clinicalcodes | | | YES |
|  | 250 AM | | | | MATURITY ONSET DIABETES (MELLITUS) | | | Clinicalcodes | | | YES |
|  | 250 AL | | | | MATURITY ONSET DIABETES(MELLITUS) NON-IN | | | Clinicalcodes | | | YES |
|  | 250 AK | | | | MATURITY ONSET DIABETES MELLITUS INSULIN | | | Clinicalcodes | | | YES |
|  | 250 AA | | | | NIDDM (NON-INSULIN DEPENDENT DIABETES) | | | Clinicalcodes | | | YES |
|  | L180600 | | | | Pre-existing diabetes mellitus; non-insulin-dependent | | | Clinicalcodes | | | YES |
|  | 66AV.00 | | | | Diabetic on insulin and oral treatment | | | Clinicalcodes | | | YES |
|  | 66A4.00 | | | | Diabetic on oral treatment | | | Clinicalcodes | | | YES |
|  | C10z100 | | | | Diabetes mellitus; adult onset; + unspecified complication | | | Clinicalcodes | | | YES |
|  | 66AH000 | | | | Conversion to insulin | | | Clinicalcodes | | | YES |
|  | C10FJ00 | | | | Insulin treated Type 2 diabetes mellitus | | | Clinicalcodes | | | YES |
|  | 66Ao.00 | | | | Diabetes type 2 review | | | Clinicalcodes | | | YES |
|  | C10FJ11 | | | | Insulin treated Type II diabetes mellitus | | | Clinicalcodes | | | YES |
|  | C10FQ00 | | | | Type 2 diabetes mellitus with exudative maculopathy | | | Clinicalcodes | | | YES |
|  | C10FP00 | | | | Type 2 diabetes mellitus with ketoacidotic coma | | | Clinicalcodes | | | YES |
|  | C10FN11 | | | | Type II diabetes mellitus with ketoacidosis | | | Clinicalcodes | | | YES |
|  | C10FN00 | | | | Type 2 diabetes mellitus with ketoacidosis | | | Clinicalcodes | | | YES |
|  | C10FM11 | | | | Type II diabetes mellitus with persistent microalbuminuria | | | Clinicalcodes | | | YES |
|  | C10FM00 | | | | Type 2 diabetes mellitus with persistent microalbuminuria | | | Clinicalcodes | | | YES |
|  | C10FL11 | | | | Type II diabetes mellitus with persistent proteinuria | | | Clinicalcodes | | | YES |
|  | C10FL00 | | | | Type 2 diabetes mellitus with persistent proteinuria | | | Clinicalcodes | | | YES |
|  | C10FK11 | | | | Hyperosmolar non-ketotic state in type II diabetes mellitus | | | Clinicalcodes | | | YES |
|  | C10F500 | | | | Type 2 diabetes mellitus with gangrene | | | Clinicalcodes | | | YES |
|  | C10y100 | | | | Diabetes mellitus; adult; + other specified manifestation | | | Clinicalcodes | | | YES |
|  | C10F600 | | | | Type 2 diabetes mellitus with retinopathy | | | Clinicalcodes | | | YES |
|  | 679R.00 | | | | Patient offered diabetes structured education programme | | | Clinicalcodes | | | YES |
|  | 889A.00 | | | Diab mellit insulin-glucose infus acute myocardial infarct | | | | | Clinicalcodes | | YES |
|  | 8CA4100 | | | Pt advised re diabetic diet | | | | | Clinicalcodes | | YES |
|  | 8CP2.00 | | | Transition of diabetes care options discussed | | | | | Clinicalcodes | | YES |
|  | 8CR2.00 | | | Diabetes clinical management plan | | | | | Clinicalcodes | | YES |
|  | 8CS0.00 | | | Diabetes care plan agreed | | | | | Clinicalcodes | | YES |
|  | 8H4e.00 | | | Referral to diabetes special interest general practitioner | | | | | Clinicalcodes | | YES |
|  | 8Hj3.00 | | | Referral to DAFNE diabetes structured education programme | | | | | Clinicalcodes | | YES |
|  | 6761 | | | Diabetic pre-pregnancy counselling | | | | | Clinicalcodes | | YES |
|  | 8I3k.00 | | | Insulin therapy declined | | | | | Clinicalcodes | | YES |
|  | 8I57.00 | | | Patient held diabetic record declined | | | | | Clinicalcodes | | YES |
|  | 9360 | | | Patient held diabetic record issued | | | | | Clinicalcodes | | YES |
|  | C10F.00 | | | Type 2 diabetes mellitus | | | | | Clinicalcodes | | YES |
|  | C10F411 | | | Type II diabetes mellitus with ulcer | | | | | Clinicalcodes | | YES |
|  | C10F400 | | | Type 2 diabetes mellitus with ulcer | | | | | Clinicalcodes | | YES |
|  | C10F311 | | | Type II diabetes mellitus with multiple complications | | | | | Clinicalcodes | | YES |
|  | C10F300 | | | Type 2 diabetes mellitus with multiple complications | | | | | Clinicalcodes | | YES |
|  | C10F211 | | | Type II diabetes mellitus with neurological complications | | | | | Clinicalcodes | | YES |
|  | C10F200 | | | Type 2 diabetes mellitus with neurological complications | | | | | Clinicalcodes | | YES |
|  | C10F111 | | | Type II diabetes mellitus with ophthalmic complications | | | | | Clinicalcodes | | YES |
|  | C10F100 | | | Type 2 diabetes mellitus with ophthalmic complications | | | | | Clinicalcodes | | YES |
|  | C10F011 | | | Type II diabetes mellitus with renal complications | | | | | Clinicalcodes | | YES |
|  | 679L.00 | | | Health education - diabetes | | | | | Clinicalcodes | | YES |
|  | C10F.11 | | | Type II diabetes mellitus | | | | | Clinicalcodes | | YES |
|  | C10F511 | | | Type II diabetes mellitus with gangrene | | | | | Clinicalcodes | | YES |
|  | C10E912 | | | Insulin dependent diabetes maturity onset | | | | | Clinicalcodes | | YES |
|  | C10D.11 | | | Maturity onset diabetes in youth type 2 | | | | | Clinicalcodes | | YES |
|  | C10D.00 | | | Diabetes mellitus autosomal dominant type 2 | | | | | Clinicalcodes | | YES |
|  | 1434 | | | H/O: diabetes mellitus | | | | | Clinicalcodes | | YES |
|  | 14P3.00 | | | H/O: insulin therapy | | | | | Clinicalcodes | | YES |
|  | 3882 | | | Diabetes well being questionnaire | | | | | Clinicalcodes | | YES |
|  | C10F000 | | | Type 2 diabetes mellitus with renal complications | | | | | Clinicalcodes | | YES |
|  | C109.11 | | | NIDDM - Non-insulin dependent diabetes mellitus | | | | | Clinicalcodes | | YES |
|  | E11.7 | | | non-insulin-dependent diabetes mellitus | | | | | Clinicalcodes | | YES |
|  | E11.8 | | | non-insulin-dependent diabetes mellitus | | | | | Clinicalcodes | | YES |
|  | E11.9 | | | non-insulin-dependent diabetes mellitus | | | | | Clinicalcodes | | YES |
|  | C104100 | | | Diabetes mellitus; adult onset; with renal manifestation | | | | | Clinicalcodes | | YES |
|  | 66AH100 | | | conversion to insulin in secondary care | | | | | Clinicalcodes | | YES |
|  | C105100 | | | Diabetes mellitus; adult onset; + ophthalmic manifestation | | | | | Clinicalcodes | | YES |
|  | C103y00 | | | Other specified diabetes mellitus with coma | | | | | Clinicalcodes | | YES |
|  | C103100 | | | Diabetes mellitus; adult onset; with ketoacidotic coma | | | | | Clinicalcodes | | YES |
|  | C109211 | | | Type II diabetes mellitus with neurological complications | | | | | Clinicalcodes | | YES |
|  | C109.00 | | | Non-insulin dependent diabetes mellitus | | | | | Clinicalcodes | | YES |
|  | E11.4 | | | non-insulin-dependent diabetes mellitus | | | | | Clinicalcodes | | YES |
|  | C109.12 | | | Type 2 diabetes mellitus | | | | | Clinicalcodes | | YES |
|  | C109.13 | | | Type II diabetes mellitus | | | | | Clinicalcodes | | YES |
|  | C109000 | | | Non-insulin-dependent diabetes mellitus with renal comps | | | | | Clinicalcodes | | YES |
|  | C109011 | | | | | Type II diabetes mellitus with renal complications | | | Clinicalcodes | | YES |
|  | C109012 | | | | | Type 2 diabetes mellitus with renal complications | | | Clinicalcodes | | YES |
|  | C109100 | | | | | Non-insulin-dependent diabetes mellitus with ophthalm comps | | | Clinicalcodes | | YES |
|  | C10P111 | | | | | Type 2 diabetes mellitus in remission | | | Clinicalcodes | | YES |
|  | C109112 | | | | | Type 2 diabetes mellitus with ophthalmic complications | | | Clinicalcodes | | YES |
|  | C102100 | | | | | Diabetes mellitus; adult onset; with hyperosmolar coma | | | Clinicalcodes | | YES |
|  | C107100 | | | | | Diabetes mellitus; adult; + peripheral circulatory disorder | | | Clinicalcodes | | YES |
|  | 8I2P.00 | | | | | sulphonylureas contraindicated | | | Clinicalcodes | | YES |
|  | C10P100 | | | | | Type II diabetes mellitus in remission | | | Clinicalcodes | | YES |
|  | C10FP11 | | | | | Type II diabetes mellitus with ketoacidotic coma | | | Clinicalcodes | | YES |
|  | ZV6DB00 | | | | | [v]admitted for conversion to insulin | | | Clinicalcodes | | YES |
|  | C106100 | | | | | Diabetes mellitus; adult onset; + neurological manifestation | | | Clinicalcodes | | YES |
|  | E11.6 | | | | | non-insulin-dependent diabetes mellitus | | | Clinicalcodes | | YES |
|  | E11.5 | | | | | non-insulin-dependent diabetes mellitus | | | Clinicalcodes | | YES |
|  | E11 | | | | | non-insulin-dependent diabetes mellitus | | | Clinicalcodes | | YES |
|  | E11.0 | | | | | non-insulin-dependent diabetes mellitus | | | Clinicalcodes | | YES |
|  | E11.1 | | | | | non-insulin-dependent diabetes mellitus | | | Clinicalcodes | | YES |
|  | E11.2 | | | | | non-insulin-dependent diabetes mellitus | | | Clinicalcodes | | YES |
|  | E11.3 | | | | | non-insulin-dependent diabetes mellitus | | | Clinicalcodes | | YES |
|  | C109200 | | | | | Non-insulin-dependent diabetes mellitus with neuro comps | | | Clinicalcodes | | YES |
|  | C109912 | | | | | Type 2 diabetes mellitus without complication | | | Clinicalcodes | | YES |
|  | C109212 | | | | | Type 2 diabetes mellitus with neurological complications | | | Clinicalcodes | | YES |
|  | C109111 | | | | | Type II diabetes mellitus with ophthalmic complications | | | Clinicalcodes | | YES |
|  | C109712 | | | | | Type 2 diabetes mellitus - poor control | | | Clinicalcodes | | YES |
|  | C100112 | | | | | Non-insulin dependent diabetes mellitus | | | Clinicalcodes | | YES |
|  | C100111 | | | | | Maturity onset diabetes | | | Clinicalcodes | | YES |
|  | C100100 | | | | | Diabetes mellitus; adult onset; no mention of complication | | | Clinicalcodes | | YES |
|  | C109D11 | | | | | Type II diabetes mellitus with hypoglycaemic coma | | | Clinicalcodes | | YES |
|  | C109612 | | | | | Type 2 diabetes mellitus with retinopathy | | | Clinicalcodes | | YES |
|  | C109911 | | | | | Type II diabetes mellitus without complication | | | Clinicalcodes | | YES |
|  | C109700 | | | | | Non-insulin dependent diabetes mellitus - poor control | | | Clinicalcodes | | YES |
|  | C109A00 | | | | | Non-insulin dependent diabetes mellitus with mononeuropathy | | | Clinicalcodes | | YES |
|  | C109A11 | | | | | Type II diabetes mellitus with mononeuropathy | | | Clinicalcodes | | YES |
|  | C109B00 | | | | | Non-insulin dependent diabetes mellitus with polyneuropathy | | | Clinicalcodes | | YES |
|  | C109B11 | | | | | Type II diabetes mellitus with polyneuropathy | | | Clinicalcodes | | YES |
|  | C109B12 | | | | | Type 2 diabetes mellitus with polyneuropathy | | | Clinicalcodes | | YES |
|  | C109C00 | | | | | Non-insulin dependent diabetes mellitus with nephropathy | | | Clinicalcodes | | YES |
|  | C109C11 | | | | | Type II diabetes mellitus with nephropathy | | | Clinicalcodes | | YES |
|  | C109C12 | | | | | Type 2 diabetes mellitus with nephropathy | | | Clinicalcodes | | YES |
|  | C109D00 | | | | | Non-insulin dependent diabetes mellitus with hypoglyca coma | | | Clinicalcodes | | YES |
|  | C109900 | | | | | Non-insulin-dependent diabetes mellitus without complication | | | Clinicalcodes | | YES |
|  | C109600 | | | | | Non-insulin-dependent diabetes mellitus with retinopathy | | | Clinicalcodes | | YES |
|  | C109300 | | | | | Non-insulin-dependent diabetes mellitus with multiple comps | | | Clinicalcodes | | YES |
|  | C109312 | | | | | Type 2 diabetes mellitus with multiple complications | | | Clinicalcodes | | YES |
|  | C109400 | | | | | Non-insulin dependent diabetes mellitus with ulcer | | | Clinicalcodes | | YES |
|  | C109411 | | | | | Type II diabetes mellitus with ulcer | | | Clinicalcodes | | YES |
|  | | C109412 | | | | | Type 2 diabetes mellitus with ulcer | | Clinicalcodes | | YES |
|  | | C109500 | | | | | Non-insulin dependent diabetes mellitus with gangrene | | Clinicalcodes | | YES |
|  | | C109511 | | | | | Type II diabetes mellitus with gangrene | | Clinicalcodes | | YES |
|  | | C109711 | | | | | Type II diabetes mellitus - poor control | | Clinicalcodes | | YES |
|  | | C107400 | | | | | NIDDM with peripheral circulatory disorder | | Clinicalcodes | | YES |
|  | | 66o2.00 | | | | | Diabetic on non-insulin injectable medication | | Clinicalcodes | | YES |
|  | | C101100 | | | | | Diabetes mellitus; adult onset; with ketoacidosis | | Clinicalcodes | | YES |
|  | | 66o5.00 | | | | | Diabetic on oral treatment and glucagon-like pepti | | Clinicalcodes | | YES |
|  | | C109611 | | | | | Type II diabetes mellitus with retinopathy | | Clinicalcodes | | YES |
|  | | C101000 | | | | | Diabetes mellitus; juvenile type; with ketoacidosis | | Clinicalcodes | | YES |
|  | | C109512 | | | | | Type 2 diabetes mellitus with gangrene | | Clinicalcodes | | YES |
|  | | 66A3.00 | | | | | Diabetic on diet only | | Clinicalcodes | | YES |

### B Data Extraction

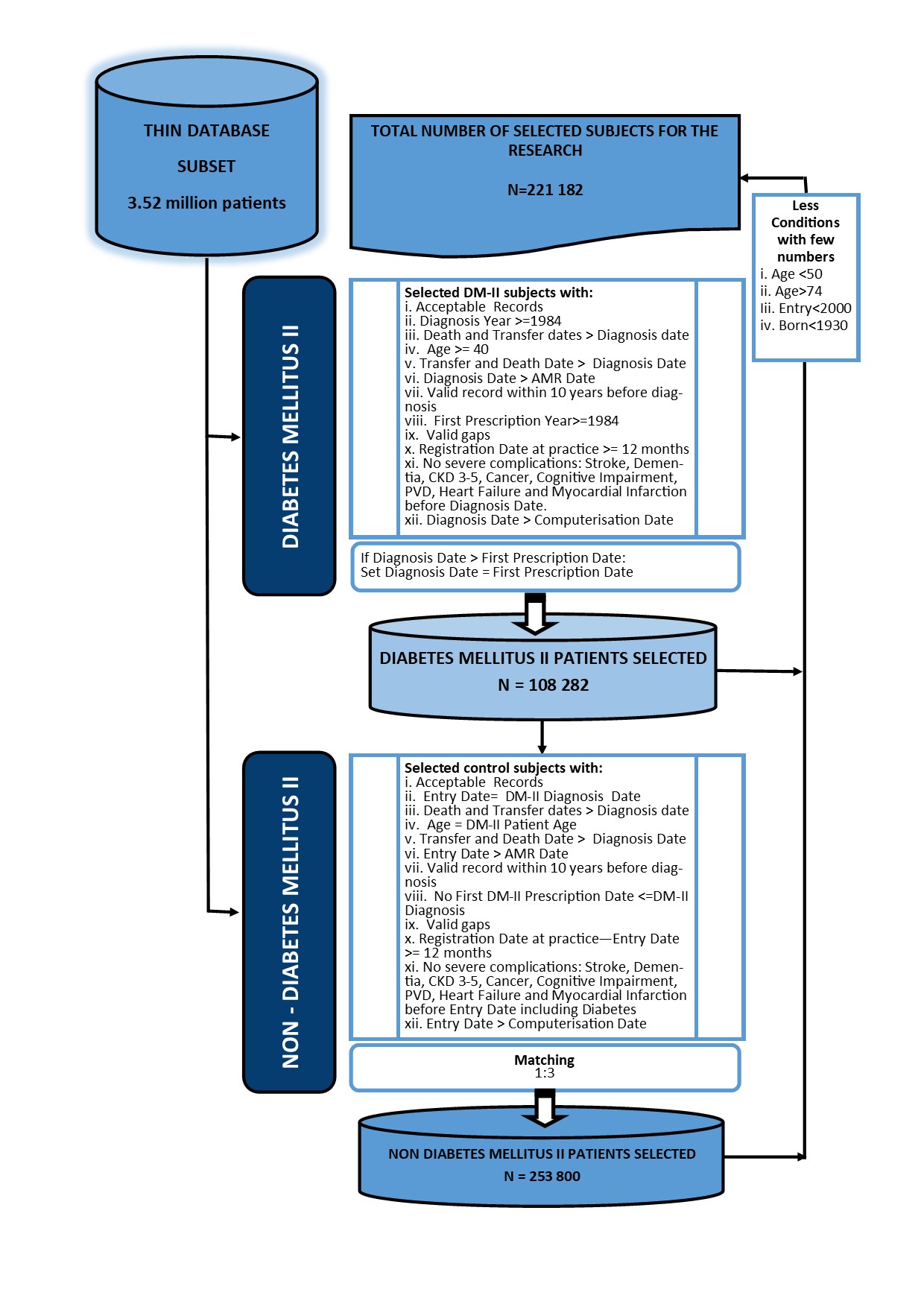

Figure S1: Data Extraction Flow Chart

### C Variable Description

Body mass index (BMI), hypertension (HTN) and hypercholesterolemia (HCL) were transformed as follows.

#### C.1 BMI

As per advice from [18], body mass index (BMI) values which were less than 13 and greater than 100 were considered to be missing values. These were handled through the multiple imputation process. Underweight BMI values were coded as normal weight as there were very few underweight individuals in this category.

#### C.2 HTN

Individuals who had a prior diagnosis of HTN or had a BP greater than 140 and had an antihypertensive drug prescription were classified as having *treated HTN* while those with no antihypertensive drug prescription were classified as having *untreated HTN*. Individuals with no diagnosis and no antihypertensive prescription at baseline were classified as having no known HTN.

#### C.3 HCL

As with the HTN, the same was done for HCL except that total cholesterol greater than 5 was used in addition for identification of HCL.

#### C.4 TDI

The Townsend Deprivation Index (TDI) is one of the several indices used to measure deprivation in the UK. Other indices are the Index of Multiple Deprivation and the Mosaic Index. The TDI is computed as a weighted index from four indicators, which are Unemployment, Non-car ownership, Non-house ownership and Overcrowding. The TDI used by the THIN database were based on the 2001 Census data. The indicators were computed as follows:

1. $unemployment= \frac{number of unemployed active labour force}{total number of active labour force} \cdot100$
2. $non-car ownership= \frac{number of households with no car}{total number of hpuseholds} \cdot100$
3. $non-house ownership= \frac{number of households tenant occupied}{total number of households} \cdot100$
4. $overcrowding= \frac{number of households overcrowded}{total number of households} \cdot100$

Their standard Z-scores were calculated, weighted and summed up to give the TDI score. THIN used equally weighted Z-scores which were based on Office of National Statistics (ONS) 2001 census method. Unemployment and overcrowding indicators were first log transformed before their Z-scores were calculated to reduce skewness. The scores were then grouped into TDI quintiles from 1 (least deprived) to 5 (most deprived). Due to the fact that the standard Z-scores are centred on the mean zero, an area with TDI score greater than zero was categorised as deprived whereas affluent areas had a score less than zero.

### D Begun et. al (2019) Model

The hazard functions for the family of Cox models can be generalised as $\lambda_{1}\left( t \right)=\lambda_{0}\left( t,\alpha,\gamma,g \right)\cdot\exp\left( \boldsymbol{\beta}^{T}\boldsymbol{X} \right)$, where α and γ are the scale and shape parameters of the baseline hazard function and $g=g(\boldsymbol{\beta}_{time-variant},\boldsymbol{X}_{time-variant})$ a function that controls for time-variant variables in X. When g = 1, the simple Cox regression is returned. [26] defined $g=exp(\boldsymbol{\beta}_{time-variant}^{T}\boldsymbol{X}_{time-variant})$. This study applied this model using the Gompertz hazard function and gamma(θ, θ) as the frailty distribution. The model has four main input which include: (1) shape parameter formula, (2) scale (or HR) parameter formula, (3) name of the hazard function and (4) frailty/cluster variable, and returns. The Gompertz shape $(\gamma)$ and scale $(\alpha)$ with shape reparametrised as υ∗ = $\gamma\cdot exp(\boldsymbol{\beta}_{time-variant}^{T}\boldsymbol{X}_{time-variant})$. The cumulative hazard function (c.h.f) is then defined as

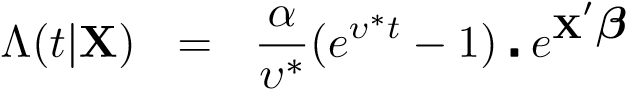
*scale*

and its cumulative survival function with a frailty from Gamma(1*/θ,*1*/θ*) specified as

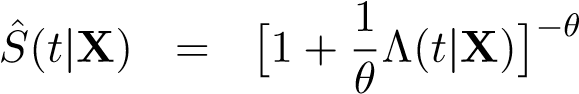
*.*

### E Additional Table

Table S2: Gompertz Cox Model with Frailty (Complete Case)

|  | | | Adjusted Gompertz Distribution Estimates | | | | | |  |
| --- | --- | --- | --- | --- | --- | --- | --- | --- | --- |
| Parameter | | | Estimate 95% CI | | p-value | | | |  |
| Sample size | | | 154 045 | |  | | | |  |
| Number of deaths | | | 19 604 | |  | | | |  |
| 1000a (scale) | 6.64 6.07 - 7.25 | | | | *<* 1*e*-16 | | | |  |
| 100b (shape) | 8.1 7.31 - 8.98 | | | | *<* 1*e*-16 | | | |  |
| Exponentiated adjusted estimates of time-variant effects | | | | | | | | | |
|  | | Shape | | | | | Scale | | |
|  | Estimate | | | 95% CI | p-value | Estimate | | 95% CI | p-value |
| Birth Cohort  1930-1939 | 1 | | |  |  | 1 | |  |  |
| 1940-1949 | 0.69 | | | 0.62 - 0.77 | *<* 1*e*-16 | 0.73 | | 0.68 - 0.79 | *<* 1*e*-16 |
| 1950-1960 | 0.89 | | | 0.76 - 1.03 | 0.12 | 0.45 | | 0.4 - 0.5 | *<* 1*e*-16 |
| AF | 1.36 | | | 1.24 - 1.49 | *<* 1*e*-16 | 0.83 | | 0.76 - 0.9 | *<* 1*e*-16 |
| HTN  None | 1 | | |  |  | 1 | |  |  |
| Treated | 1.37 | | | 1.24 - 1.52 | *<* 1*e*-16 | 0.81 | | 0.76 - 0.86 | *<* 1*e*-16 |
| Untreated | 0.89 | | | 0.77 - 1.03 | 0.11 | 1.48 | | 1.38 - 1.58 | *<* 1*e*-16 |

Frailty Estimate

Variance (*σ*^2^)	0.12	0.1 - 0.14 *<* 1*e*-16

Goodness of Fit

Concordance (*σ*)	0.74 (0.002)

Log Likelihood	-96410.95

AIC	192907.9

### F Additional Figures

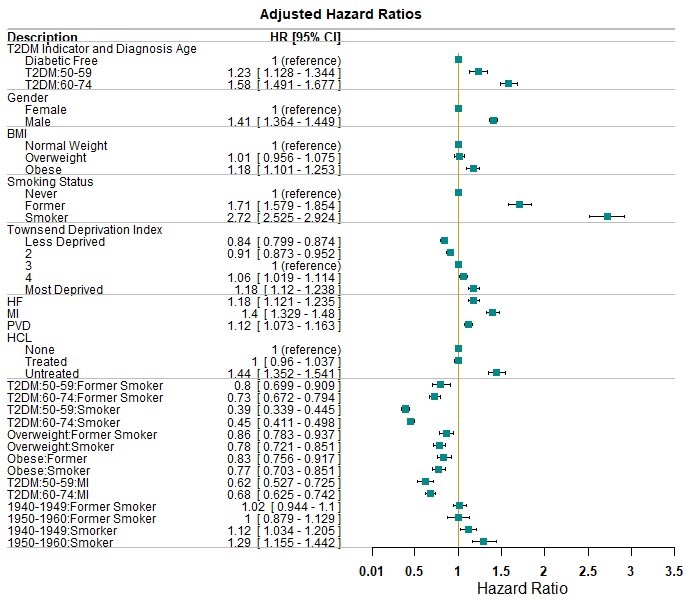

Figure S2: Hazard Ratios for the complete case model

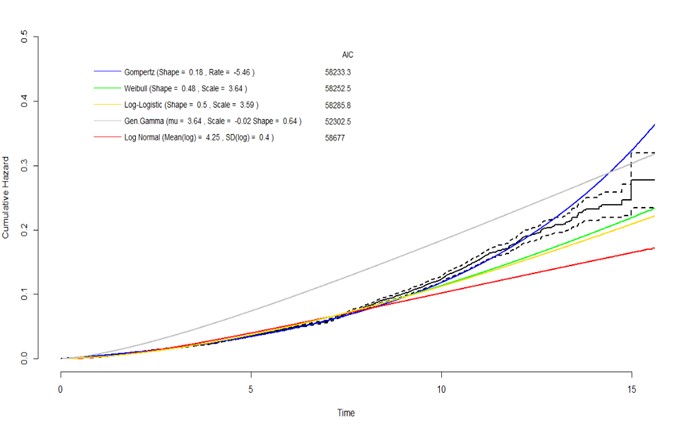

Figure S3: Hazard function distribution
